# Sub-Threshold Stochastic Vibration Improves Fingertip Tactile Sensitivity in Stroke Survivors and Healthy Adults, with Source-Localized EEG Evidence for Cortical Network Reorganization in Healthy Adults

**DOI:** 10.64898/2026.11.10.26367694

**Authors:** Moeez Ashfaque, Usman Ghani, Sandeep Bhagwat, Simon F. Farmer, Imran Khan Niazi, Amit N. Pujari

## Abstract

Cutaneous sensation is vital for daily function and safety, yet it commonly declines with ageing and after neurological injury, with stroke survivors particularly affected. This study investigates whether sub-threshold stochastic (random) vibrations can enhance light-touch (cutaneous) sensation in healthy adults and stroke survivors and characterizes the associated changes in cortical functional connectivity (FC). Nineteen healthy participants and five stroke survivors took part. A custom device delivered sinusoidal and stochastic vibrations to the volar surface of the right middle fingertip. For each participant we first determined the detection threshold for stochastic vibration alone, then the threshold for a 30 Hz sinusoidal vibration, and then the threshold for that sinusoidal vibration combined with imperceptible stochastic noise fixed at 80% of the stochastic threshold; in ten healthy participants a super-threshold combination (120% of the stochastic threshold) was also tested. High-density (64-channel) electroencephalography (EEG) was recorded throughout for the healthy participants. Adding sub-threshold stochastic noise lowered the sinusoidal detection threshold in every participant tested and produced consistent reductions in all five stroke survivors; expressed as gains in sensitivity, individual improvements reached up to 19 times in healthy participants and up to 4 times in stroke survivors. Source-localized EEG showed that sub-threshold stochastic vibration significantly increased FC, most extensively in the alpha band, with prominent involvement of frontal regions (superior and rostral middle frontal cortices and anterior cingulate; cluster-based permutation p < 0.05), consistent with a noise-driven modulation of sensory and attentional networks. Super-threshold noise produced weaker, predominantly beta-band changes, in line with signal masking rather than resonance. These findings indicate that sub-threshold stochastic vibration lowers light-touch detection thresholds and reorganizes cortical connectivity in both healthy and stroke populations, and they support prioritizing cutaneous sensation in stochastic-resonance–based neurorehabilitation for stroke survivors.

## I. Introduction

Cutaneous sensation, the perception of touch, temperature, and pain, underpins safe and effective interaction with the environment and is essential to everyday hand function [8]. These sensory capabilities frequently deteriorate with ageing and, more abruptly, after neurological injury such as stroke [1], [9]. Depending on the assessment and the time since injury, discriminative or light-touch sensory loss in the hand has been reported in roughly 35–55% of stroke survivors, and discriminative loss has been observed in almost half of chronic and up to about 85% of acute stroke survivors [2], [23]. Such deficits impair dexterity, degrade grip-force regulation, increase injury risk, and reduce quality of life [5], [23]. Because they arise from disruption of the somatosensory pathways that encode and relay tactile information [1], there is a pressing need for rehabilitation strategies that target the neurophysiological basis of the impairment rather than its symptoms alone.

Conventional sensory-rehabilitation and pharmacological approaches have had limited success in restoring the underlying sensory processing. Pharmacological agents may offer symptomatic relief but do not reliably drive the neural reorganization required for lasting recovery [19], [20], and more invasive options such as peripheral nerve stimulation carry risks that limit their routine clinical use [21]. These limitations have motivated interest in non-invasive neuromodulation, and in particular in stochastic resonance (SR), a stimulation phenomenon in which adding an optimal level of random noise to a weak, (un-detectable) sub-threshold signal improves its detection and transmission in a non-linear system [3], [4].

In the somatosensory domain, sub-threshold mechanical noise has repeatedly been shown to enhance tactile perception. Collins and colleagues first demonstrated that low-level sub-threshold vibrotactile noise improves tactile detection in healthy skin, whereas supra-threshold noise degrades it by masking the signal [4]. Liu et al. subsequently showed that adding sub-threshold mechanical noise significantly lowered vibrotactile detection thresholds at the fingertip in older adults, in patients with diabetic neuropathy, and in four of five stroke patients, establishing the clinical relevance of SR for populations with sensory loss [5]. More recently, Enders et al. reported that even remotely applied sub-threshold vibrotactile noise, i.e., delivered at the wrist or dorsal hand rather instead to the fingertip, instantaneously improved fingertip light-touch sensation in stroke survivors, independent of noise location and intensity. The authors attributed this effect to increased excitability and synchronization within the central nervous system via inter-neuronal connections [23]. The optimal noise intensity, however, remains debated: levels from approximately 50% to 90% of the sensory threshold have been reported as effective, with no consensus for stroke specifically [5], [23], motivating the sub-threshold (80%) and supra-threshold (120%) levels examined here.

Despite this consistent behavioral evidence, the cortical mechanisms that accompany noise-enhanced tactile perception remain poorly characterized. Prior work has largely inferred a central contribution indirectly, from psychophysics, evoked potentials, or the observation that remote noise can influence distant skin sites [5], [7], [23], however the frequency-specific (alpha, beta band) changes in cortical functional connectivity (FC) that underlie these perceptual gains have not been directly studied, particularly in stroke. FC, the temporal synchronization of activity between spatially separated brain regions [14], is a sensitive index of how sensory information is integrated across networks, and alpha- and beta-band synchronization in particular are implicated in attention, sensory processing, and sensorimotor integration [15], [18], [22]. Characterizing how sub-threshold stochastic vibration reshapes FC would therefore provide a mechanistic bridge between the peripheral stimulus and the perceptual outcome.

Therefore, this study combines quantitative psychophysics with source-localized, high-density EEG to answer two questions: (1) how sub-threshold stochastic vibration modulates cortical FC during tactile processing, and (2) whether these neural changes accompany measurable improvements in tactile detection thresholds. Using a within-participant design, we compared pure 30 Hz sinusoidal stimulation with sinusoidal stimulation combined with sub-threshold (80%) or supra-threshold (120%) stochastic noise, in healthy adults and stroke survivors. We hypothesized that (1) sub-threshold stochastic vibration would lower tactile detection thresholds—that is, increase sensitivity, relative to sinusoidal stimulation alone in both groups; (2) these perceptual improvements would be accompanied by increased cortical FC, predominantly in the alpha and beta bands; and (3) supra-threshold noise, being itself perceptible, would not confer a sub-threshold–type benefit and would instead be consistent with signal masking.

By linking psychophysical thresholds to source-localized FC, this work aims to establish a neurophysiological rationale for SR-based sensory rehabilitation after stroke. The remainder of the paper describes the methods (Section II), results (Section III), discussion (Section IV), and conclusions and future directions (Section V).

## II. Methods

### A. Participants

Nineteen healthy adults and five chronic stroke survivors participated. The stroke participants were recruited in Solapur, India. Healthy-participant inclusion criteria were: age 18 years or over, no history of neurological disorder, and no use of neuro-inhibitory medication in the preceding month. Stroke-participant inclusion criteria were: age 18 years or over; stroke more than six months previously; impaired light touch on two-point discrimination testing; ability to place and maintain the hand on the device plate; and absence of cognitive or severe language impairment that would preclude reliable psychophysical responding. Exclusion criteria were: an implanted electronic device such as a cardiac pacemaker; epilepsy or seizure history; a history of upper-extremity orthopedic conditions; and metalwork in the forearm. The stroke participants mean age was 60.66 ± 8.44 years; 3 male; mean time since stroke 20.75 years. All stroke participants had impaired light touch (monofilament testing: average 4.8 ± 1.0 in the middle digit of the affected hand vs 3.0 ± 0.3 in the unaffected hand).

The nineteen healthy (mean age 26.5 ± 6.84; 6 female) participants were recruited at the University of Hertfordshire (UH), UK. The stroke participants and the first healthy batch (n=9) completed vibration modes (a), (b), and (c); the healthy batch additionally underwent EEG recording. The second healthy batch (n=10) additionally completed mode (d), the super-threshold condition, allowing assessment of a super-threshold stochastic vibration. Experiments were conducted at the site of recruitment (UK or India) using a shared, harmonised protocol. Two researchers (MA and AP) conducted the experiments: MA and AP jointly in the UK, and AP alone in India. The Ethical Review Committee of the University of Hertfordshire approved the study (Reference: aSPECS/SF/UH/04703(1)). All procedures performed involving human participants followed the ethical standards of the institutional review committee and adhered to the declaration of Helsinki. An information sheet was given to the participants, and all the participants signed a ‘written consent form’ in presence of the researcher before the experiment.

### B. Procedures

#### Apparatus and Positioning

Vibrotactile stimuli were delivered to the volar surface of the right middle fingertip using a custom research apparatus developed at the University of Hertfordshire. The participant sat in a supported seated position, their affected hand was placed palm down on the device plate, on a platform that held the forearm and hand in a fixed position with a foam pad resting on top for comfort and to help maintain the position during testing, with a vibration actuator sited beneath the volar surface of the middle fingertip. The device generated four experimental modes of vibration profiles, i.e. stimuli, described below.

#### Stimuli

Two classes of vibration were delivered at a single fingertip contact site, alone or superimposed: (i) a deterministic sinusoidal vibration at 30 Hz (Fig 1, upper trace), and (ii) a zero-mean random (stochastic) vibration (Fig 1, lower trace). Four experimental modes were used: (a) stochastic vibration alone; (b) 30 Hz sinusoidal vibration alone; (c) sinusoidal vibration superimposed with sub-threshold stochastic vibration, fixed at 80% of the participant’s individually determined stochastic detection threshold; and (d) sinusoidal vibration superimposed with supra-threshold stochastic vibration at 120% of that threshold. Each stimulus presentation lasted 1 s. Stimulus amplitude was adjustable in discrete, monotonically increasing steps spanning each participant’s detection threshold; amplitudes are consequently reported throughout in arbitrary device units rather than absolute physical magnitudes. Representative sinusoidal and stochastic stimulus profiles are shown in Fig. 1.

**Figure 1.**
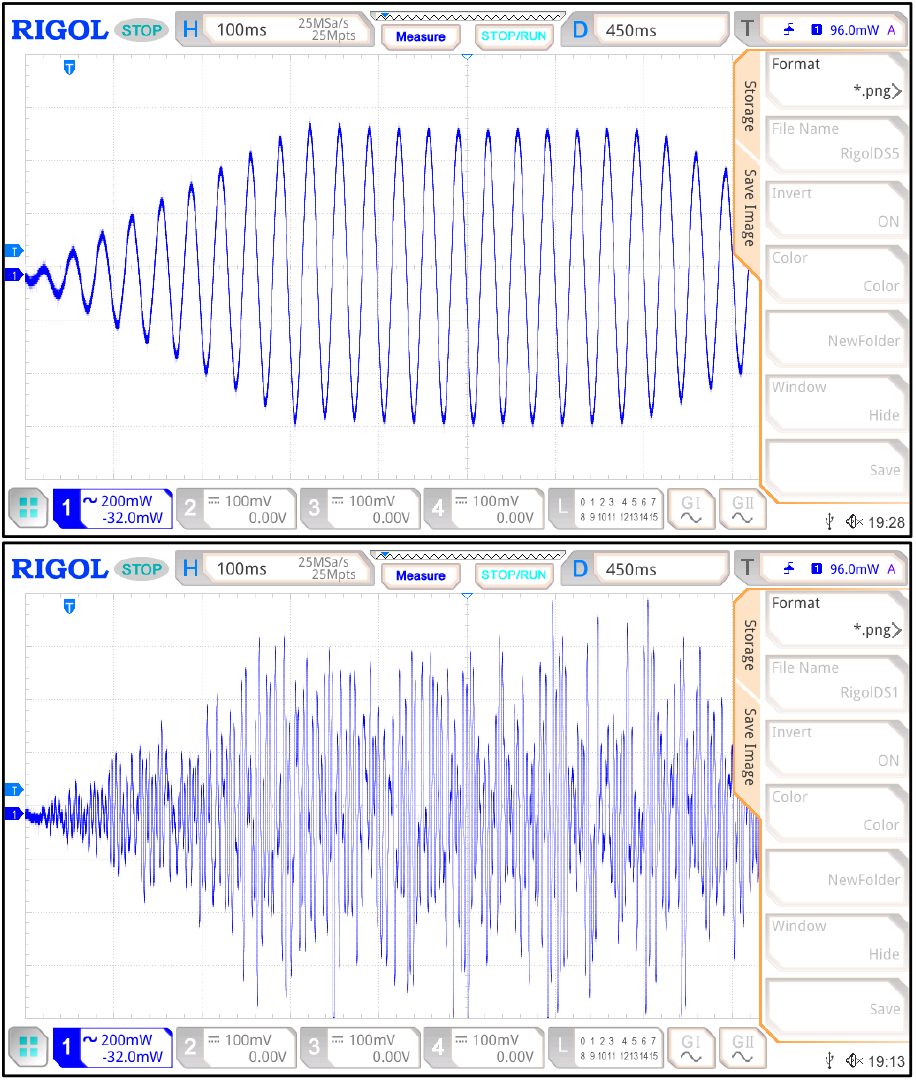
Representative samples of 30 Hz vibration stimulus (upper trace) and the stochastic noise (lower trace), oscilloscope recording.

#### Protocol

Participants rested the right hand on the device and kept it still throughout. Stimuli were delivered through the device, and participants signalled detection with the left index finger. For healthy participants, EEG was recorded continuously; to limit ocular and movement artefact, participants were asked to minimise head and eye movement and to wait 5 s after detecting a stimulus before signalling.

Testing began by determining each participant’s minimum detectable amplitude of stochastic vibration alone, i.e. the stochastic (noise) detection threshold. A sub-threshold level was then set at 80% of this value (the nearest achievable level was used where device discretization prevented an exact setting), yielding an imperceptible noise. Participants were then tested, in randomised order, on mode (b) sinusoidal vibration alone, and mode (c) sinusoidal vibration summed with the 80% sub-threshold stochastic noise. The minimum detectable amplitude was determined for each condition; these were designated the sinusoidal (normal) detection threshold [mode (b)] and the sub-threshold stochastic detection threshold [mode (c)] and were compared to quantify the effect of imperceptible noise on sensitivity. The same protocol was used for the stroke participants. For the ten healthy participants recruited at NZCC, an additional super-threshold condition [mode (d)] combined sinusoidal vibration with stochastic noise fixed at 120% of the stochastic threshold was tested. The resulting threshold was designated the super-threshold stochastic detection threshold.

Thresholds were located using a 4–2–1 incremental stepping algorithm, following the approach of Liu et al. [5]: amplitude was first adjusted in steps of four levels until the undetectable range was reached, then in steps of two levels, and finally in single levels to pinpoint the threshold. The procedure was repeated until a consistent value was obtained; if no definitive threshold was reached within 25 observations, the value at the 25th observation was recorded. All 25 observations were retained for each participant. Fig. 2 summarises the protocol.

**Figure 2.**
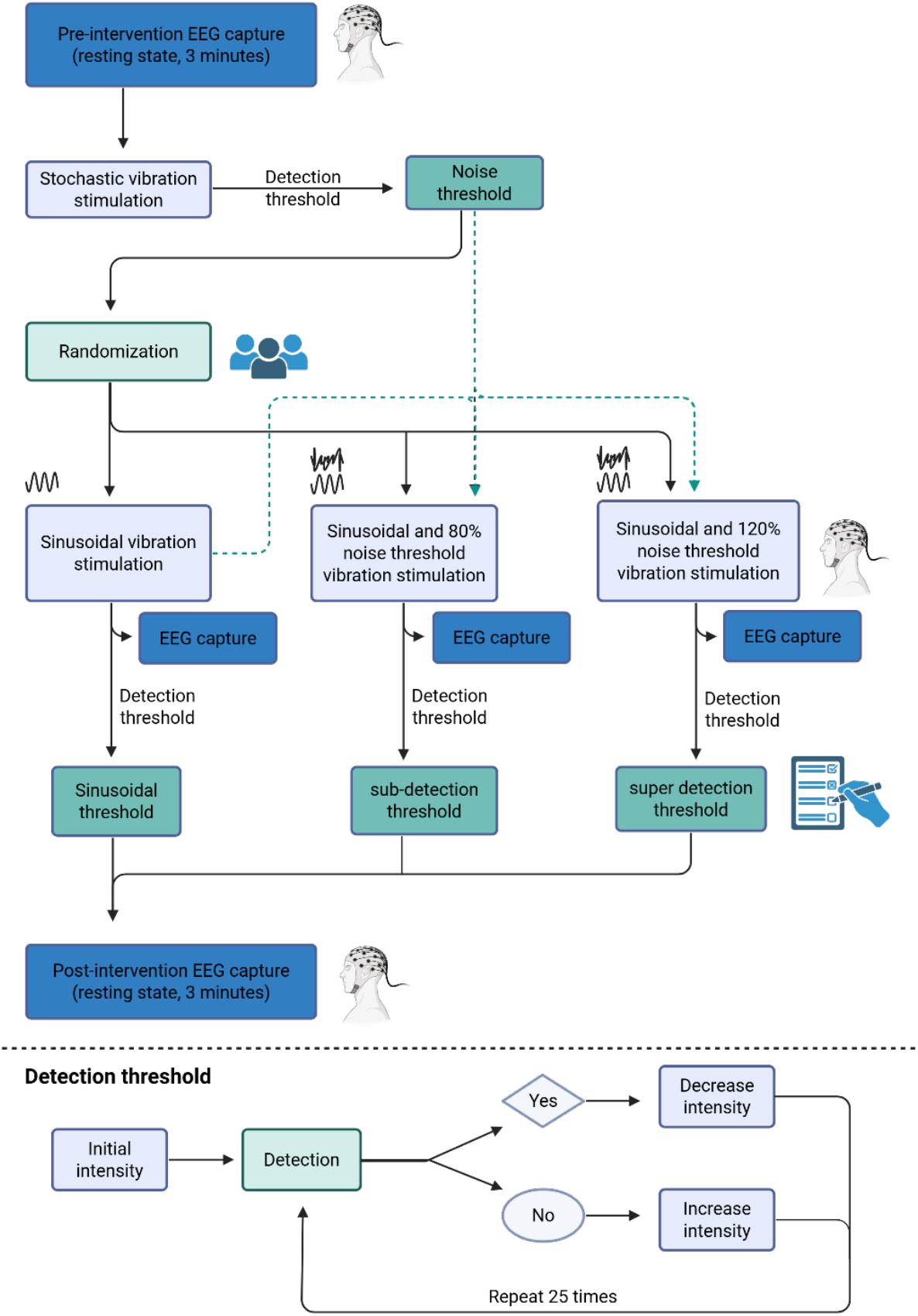
Experimental protocol. After a 3-minute resting-state EEG, the stochastic (noise) detection threshold was determined. Participants then completed, in randomized order, mode (b) sinusoidal vibration alone, mode (c) sinusoidal vibration plus noise at 80% of the noise threshold, and, in the NZCC cohort only, mode (d) sinusoidal vibration plus noise at 120%. EEG was recorded during each mode and in a 3-minute post-experiment resting state. The inset shows the up/down intensity adjustment used to locate each detection threshold (maximum 25 observations).

#### Electroencephalography (EEG) Assessments

EEG was recorded from healthy participants throughout. A 3-minute eyes-open resting-state recording was obtained before the vibration tasks, followed by recordings during each of the first three vibration modes. For the ten NZCC participants, an additional recording was obtained during mode (d) together with a 3-minute post-experiment resting-state recording. Participants were asked to keep the body, head, and eyes still to limit artefact. Data were acquired with a 64-channel cap and a TMSi REFA amplifier at 2,048 Hz using Polybench software. Electrodes followed the international 10–20 system, with the ground at AFz and both mastoids (M1, M2) as reference; impedances were kept below 10 kΩ.

### C. Data Processing

#### Data Processing

Comparisons were performed across five conditions: (1) resting-state pre (eyes open, 3 min), (2) mode (b) sinusoidal vibration, (3) mode (c) sub-threshold combination, (4) mode (d) super-threshold combination, and (5) resting-state post (eyes open, 3 min). Two outcomes were assessed: (i) cutaneous detection thresholds across conditions, to establish the clinical effect, and (ii) EEG-based FC changes between conditions, to characterise the underlying neurophysiology. Comparisons involving all nineteen participants used the full sample; comparisons involving mode (d) used the ten NZCC participants. For the psychophysical outcome, thresholds for all applicable modes were determined for each participant and compared to quantify noise-induced changes in sensitivity. EEG preprocessing (Fig. 3) proceeded as follows: the 62 scalp electrodes (excluding the two mastoids) were re-referenced to a common average; signals were band-pass filtered at 0.5–40 Hz; artefact subspace reconstruction was applied to remove high-amplitude noise; and independent component analysis with ICLabel was used to reject artefactual components using pre-trained classifiers. These steps were performed in MATLAB using EEGLAB. The pre-processed data were then imported into Brainstorm for source estimation.

**Figure 3.**
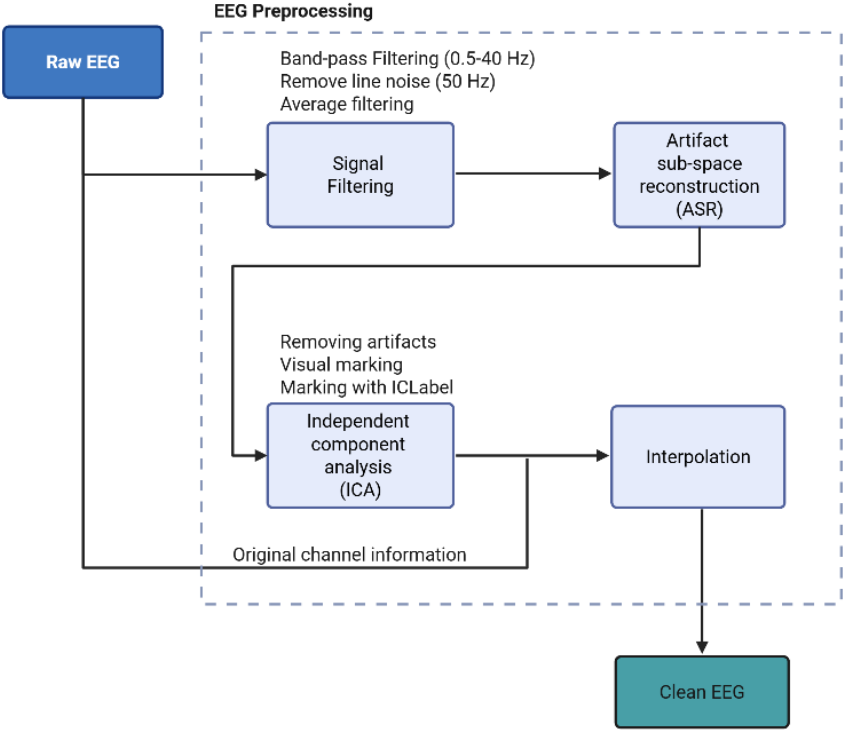
Preprocessing pipeline for EEG data processing.

#### D. Source Reconstruction and Functional Connectivity

In this study, we employed source-level EEG analysis to investigate FC changes induced by stochastic vibration. Source-level analysis allows for the localization of neural activity to specific brain regions, providing a more precise assessment of connectivity patterns compared to sensor-level approaches. This helps us identify what brain regions are associated with the vibration stimuli and what effect does it have on the functional behaviour. Our methodology involved two key steps: forward modelling and inverse modelling. Forward modelling is essential for EEG source reconstruction, as it defines the relationship between cortical sources and EEG sensor measurements. We determined the location and orientation of EEG sensors relative to cortical sources by modelling current dipole fields on a voxel grid approximating the cortical space. Dipoles were oriented perpendicular to the cortex to ensure physiological accuracy. The symmetric boundary element method (Open MEEG BEM) was used to model dipoles for all subjects [10]. A default generic head model from Brainstorm (Dec 2024) was employed, featuring 15,000 vertices and a three-layer compartment (scalp, skull, and brain). Tissue conductivities were set based on established values: scalp = 1, skull = 0.0125, and brain = 1 [11]. Electrode locations were defined using the 10-20 electrode placement system, with 64 electrodes (including references M1 and M2) positioned on the scalp.

The inverse problem in EEG source reconstruction involves estimating cortical activity from sensor measurements, which is inherently underdetermined and ill-posed due to the limited number of sensors relative to the number of potential sources. To address this, we utilized the minimum-norm estimation (MNE) method, which minimizes the overall power of estimated sources while using an identity matrix as the noise covariance matrix [12]. However, MNE tends to bias sources toward superficial cortical regions, leading to inaccuracies. To mitigate this, we applied the standardized low-resolution brain electromagnetic tomography (sLORETA) method, which normalizes current density maps and provides more accurate localization of source dipoles [13]. This source reconstruction step was performed in Brainstorm software. Fig. 4 shows the source reconstruction pipeline.

**Figure 4.**
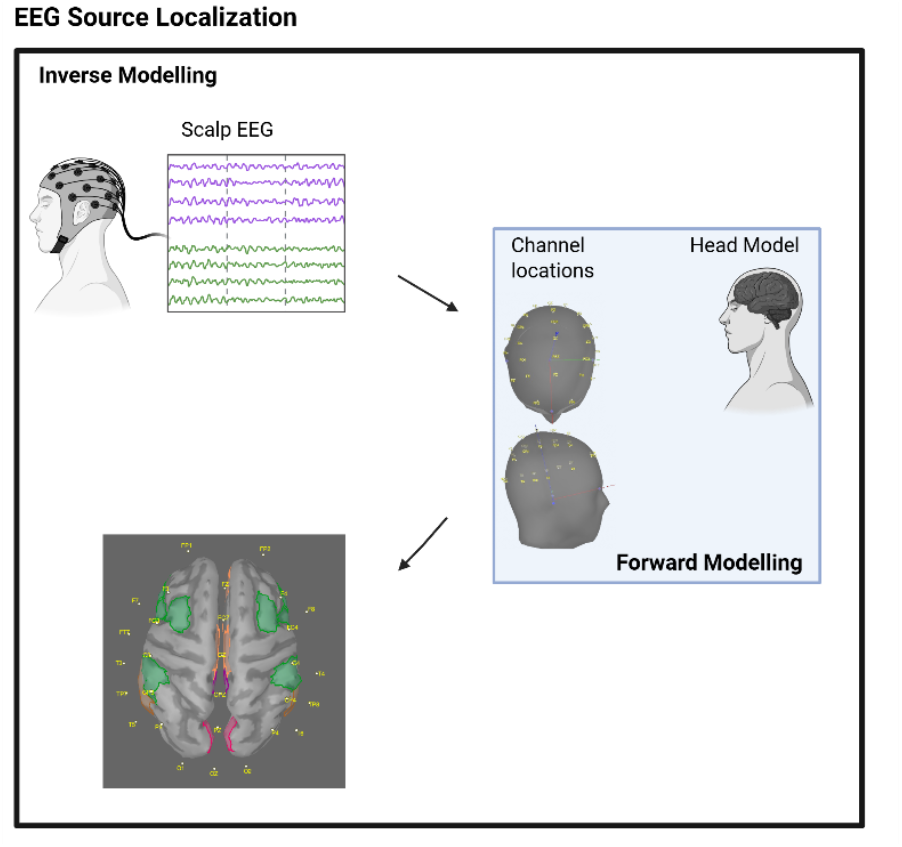
Source reconstruction pipeline. Scalp EEG is combined with a forward model (OpenMEEG BEM, three-layer generic head model, 10–20 electrode positions) and inverted using minimum-norm estimation with sLORETA normalisation to obtain cortical source activity, which is parcellated into 68 Desikan–Killiany regions.

Functional connectivity (FC) refers to the temporal correlation or synchronization of neural activity between spatially separated brain regions, reflecting their interaction and information sharing within a functional network [14]. Unlike structural connectivity, which is defined by physical pathways, FC captures dynamic relationships between brain regions, even in the absence of direct anatomical connections. It is typically quantified using metrics such as coherence, phase-locking value, or mutual information, which assess the degree of neural synchronization across regions. For functional connectivity analysis, high-resolution sources were grouped into 68 regions of interest (ROIs) based on the Desikan-Killiany atlas. Time series within each ROI were averaged to form an [ROIs × time] matrix. To prevent activity cancellation, the sign of dipoles with opposite orientations was flipped before averaging. This approach enabled robust estimation of brain activity and facilitated the assessment of connectivity patterns between distinct brain regions. Triple network (salience, central, and default mode network) were then separated out from these regions to gain a specific understanding of the FC trends.

The reconstructed EEG source signals were filtered into narrow-band signals using a 4th-order Butterworth filter to extract frequency bands of interest: alpha (7.5–12.5 Hz) and beta (12.5–30 Hz) based on [15]. To quantify FC, we employed the Phase Lag Index (PLI), a measure that assesses the asymmetry in the phase difference distribution between two signals, denoted as *x*(*t*) and *y*(*t*). PLI is particularly useful for identifying consistent phase leads or lags between signals, which are indicative of directed functional interactions.

The PLI was calculated using the following equation:

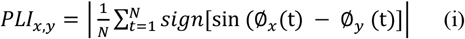

where *ϕx*(*t*) and *ϕy*(*t*) represent the instantaneous phase of signals *x*(*t*) and *y*(*t*), respectively. The phase information was derived from the Hilbert transform of the signals. The resulting phase difference can be positive, negative, or zero, depending on the relative timing of the signals. The PLI was computed over a specific time window, with *N* representing the total number of samples within that window. This process enabled us to quantify the extent of connectivity and asymmetry between the signals, providing valuable insights into the functional interactions of brain regions. The PLI was computed between every pair of source signals within each band. PLI values range from 0 (no connectivity) to 1 (maximum connectivity), capturing phase asymmetry and functional interactions. PLI values were stored in adjacency matrices (diagonal zeros, symmetric) for each frequency band and experimental modes (mode (a), mode (b), mode (c), mode (d), resting-state pre and post). Differences in PLI between experimental conditions were tested with a non-parametric cluster-based permutation test, described as follows.

## E. Statistical Analysis

To address the multiple comparison problem and control the familywise error rate, we employed a non-parametric cluster-based permutation test [16]. This test compared PLI values within adjacency matrices across experimental modes (resting-state pre vs. resting-state post, mode (b) vs. mode (c), and mode (b) vs. mode (d)) using a within-study design. The analysis was conducted using the FieldTrip toolbox (Donders Institute, Brain, Cognition, and Behavior, Nijmegen, Netherlands).

For each pairwise comparison, cluster-level statistics were computed. The largest cluster-level statistic served as the critical t-value for identifying significant clusters. A null distribution was generated by performing 5000 permutations, where data labels were randomly shuffled for each comparison. Cluster-level statistics were recalculated for each permutation to reflect the expected distribution under the null hypothesis of no connectivity differences between conditions. The p-value for each observed cluster was determined as the proportion of permutations where the cluster-level statistic exceeded the observed value. Significant clusters were classified as positive (+cluster) or negative (-cluster), indicating increased or decreased functional connectivity, respectively. This procedure identified regions with significant condition-dependent changes in connectivity while correcting for multiple comparisons.

Results were visualized using EEGNET version 1 [17], with coloured spheres representing brain regions and connecting lines depicting statistically significant functional connectivity dependencies.

## III. Results

### A. Cutaneous Sensation Thresholds

Four vibration modes were evaluated: (a) stochastic vibration alone, (b) 30 Hz sinusoidal vibration, (c) sinusoidal vibration with sub-threshold (80%) stochastic noise, and (d) sinusoidal vibration with super-threshold (120%) stochastic noise. The primary comparisons contrasted the sinusoidal threshold [mode (b)] with the sub-threshold [mode (c)] and super-threshold [mode (d)] conditions. Healthy participants completed all modes; stroke participants completed modes (a)–(c). Throughout, a lower detection threshold indicates greater sensitivity; the per-participant increase in sensitivity reported below is defined as:

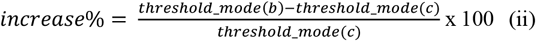

Healthy participants showed substantial inter-individual variability in baseline sensitivity but a consistent direction of effect. The stochastic threshold [mode (a)] was 0.3 units in 11 participants, 0.4 in 6, and 0.2 and 0.9 in one participant each. Sinusoidal thresholds [mode (b)] ranged from 2 to 20 units. Adding sub-threshold noise [mode (c)] lowered the threshold in every participant, to a range of 0–10 units (most between 1 and 5). Broadly, participants with a low stochastic threshold also tended to have a low sinusoidal threshold, though this was not universal (e.g., participants 1 and 3). The corresponding gains in sensitivity ranged from 60% to 1900%, with participant 13 showing the largest gain (1900%) and participants 6, 10, 16, 17, and 18 showing 100%. In mode (d), thresholds were recorded as 0 for all participants, as expected, since the 120% noise is itself supra-threshold and therefore always perceptible, so the minimum presented level already exceeded detection. Individual values are given in Table II, and Fig. 5 shows the clear reduction in mode (c) relative to mode (b) across participants.

**Figure 5.**
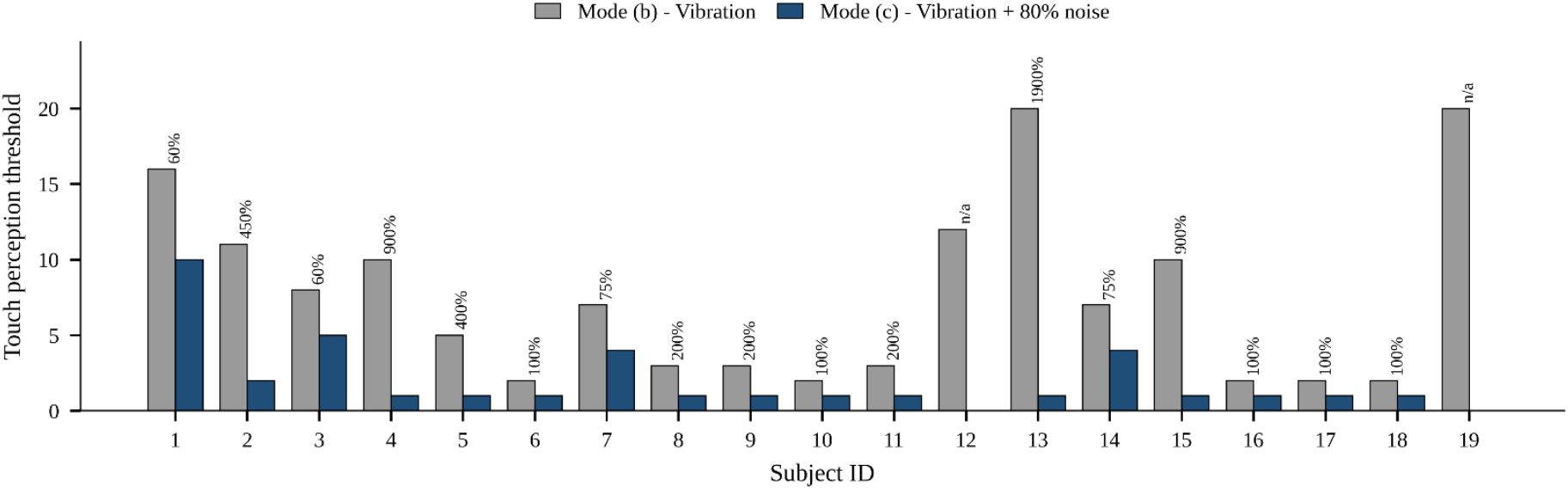
Touch perception thresholds for the nineteen healthy participants in mode (b) (30 Hz sinusoidal vibration) and mode (c) (sinusoidal vibration plus noise at 80% of the stochastic threshold). Thresholds are in arbitrary device units. Percentages above each pair give the sensitivity gain, (b − c)/c × 100%. A gain is not shown (“n/a”) for participants 12 and 19, whose mode (c) threshold was 0.

All five stroke participants showed a lower threshold in mode (c) than in mode (b)—that is, improved sensitivity with imperceptible noise—with gains ranging from 16% to 400%.

The stochastic threshold [mode (a)] was comparatively uniform (1 unit in three participants, 0 in one, and 1.8 in one). Individual values are given in Table I, and Fig. 6 illustrates the reduction in detection threshold for the noise-added condition relative to sinusoidal stimulation alone.

**Figure 6.**
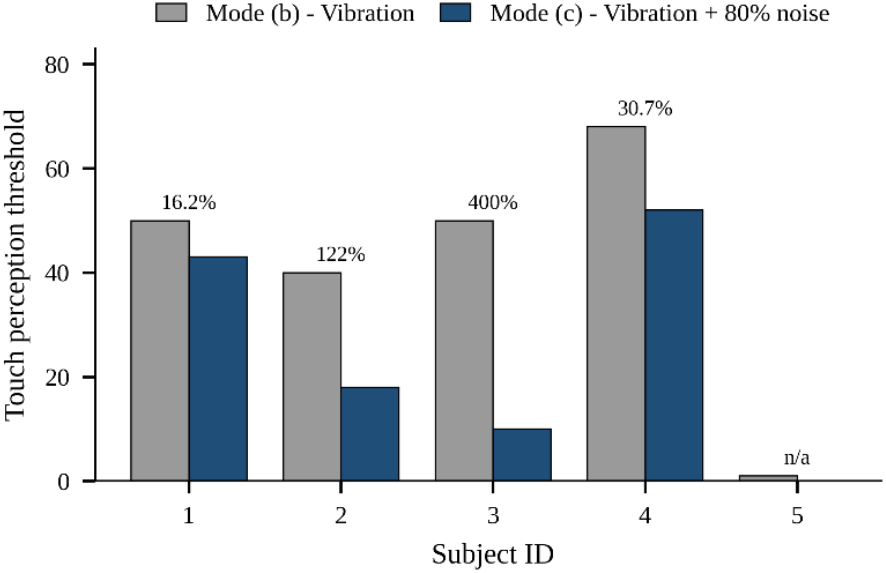
Touch perception thresholds for the five stroke participants in mode (b) (30 Hz sinusoidal vibration) and mode (c) (sinusoidal vibration plus noise at 80% of the stochastic threshold). Percentages above each pair give the sensitivity gain, (b − c)/c × 100%. The gain is not defined for participant 5, whose mode (c) threshold was 0.

**TABLE 1.**
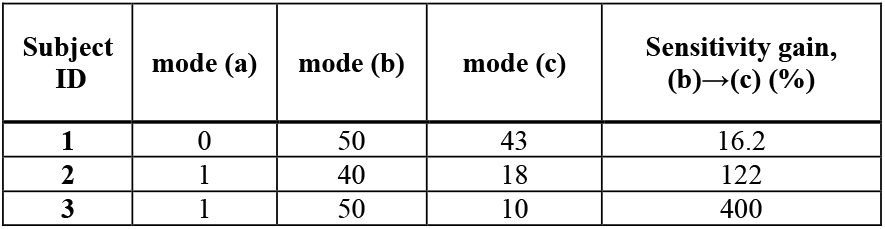
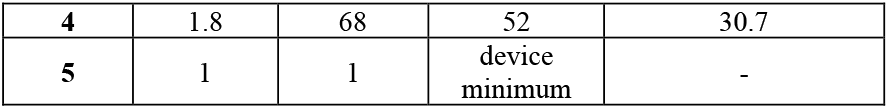
Cutaneous sensation thresholds for stroke participants: (A) stochastic threshold, (B) sinusoidal vibration threshold, and (C) sub-threshold combination. The final column reports the gain in sensitivity from mode (B) to mode (C).

### B. Functional Connectivity (FC)

Source-localised EEG from the nineteen healthy participants revealed that stochastic vibration significantly changed the cortical FC, within both the alpha and beta bands, relative to sinusoidal stimulation and between resting states, as quantified by the PLI and tested with cluster-based permutation.

For each comparison, cluster-based permutation testing identified spatially contiguous sets of connections that differed significantly between conditions, controlling for multiple comparisons. The reported statistics (Tables III and IV, for the alpha and beta bands) correspond to the significant clusters and their associated p- and effect (RHO) values. The connectivity changes were frequency- and condition-dependent. Sub-threshold stimulation [mode (b) vs. (c)] produced the most widespread alpha-band changes (24 significant region pairs) alongside far fewer beta-band changes (4 pairs); the resting-state contrast involved 17 pairs in each band; and the super-threshold contrast [mode (b) vs. (d)] produced only two alpha-band changes with a predominantly beta-band signature. Significant clusters in the sub-threshold contrast prominently involved frontal regions—including the superior and rostral middle frontal cortices, rostral anterior cingulate, and insula— consistent with modulation of sensory and attentional networks (Figs. 7 and 8). Significant networks were rendered at the cortical surface using EEGNET (version 1) [17] and custom circular-graph plots.

**Figure 7.**
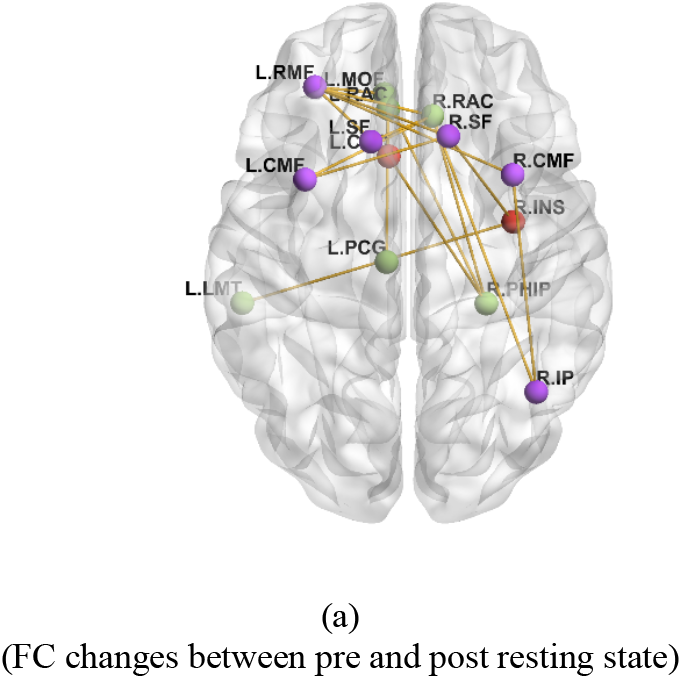
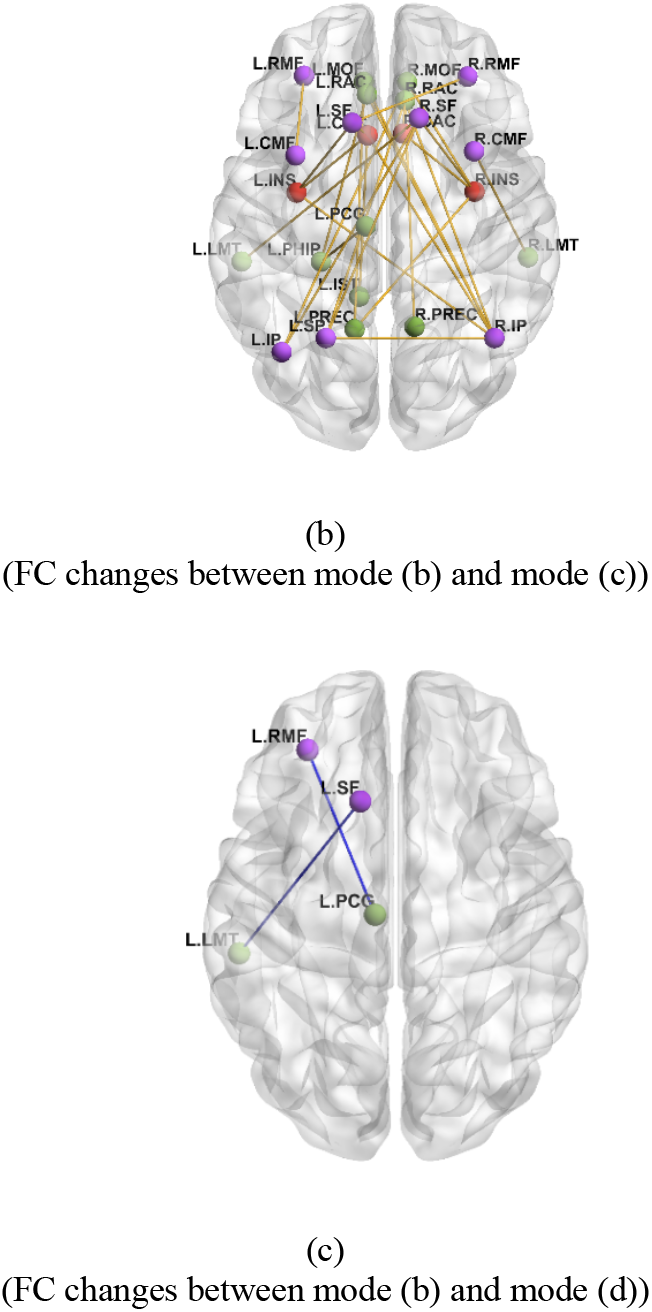
Significant changes in functional connectivity (PLI) in the alpha band between (a) resting-state pre and post, (b) mode-(b) and mode-(c), and (c) mode-(b) and mode-(d). Spheres are cortical regions, coloured by network (1–3); lines are connections with significant differences (cluster-based permutation test, p < 0.05). Orange coloured lines indicates increase whereas blue decrease in the activity. Nodes are colored as: purple is CEN, green is DMN, and red is the Salience Network.

**Figure 8.**
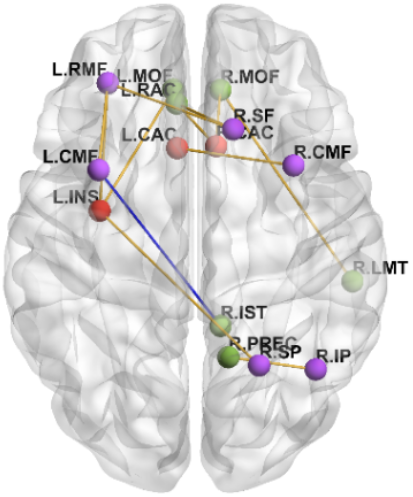
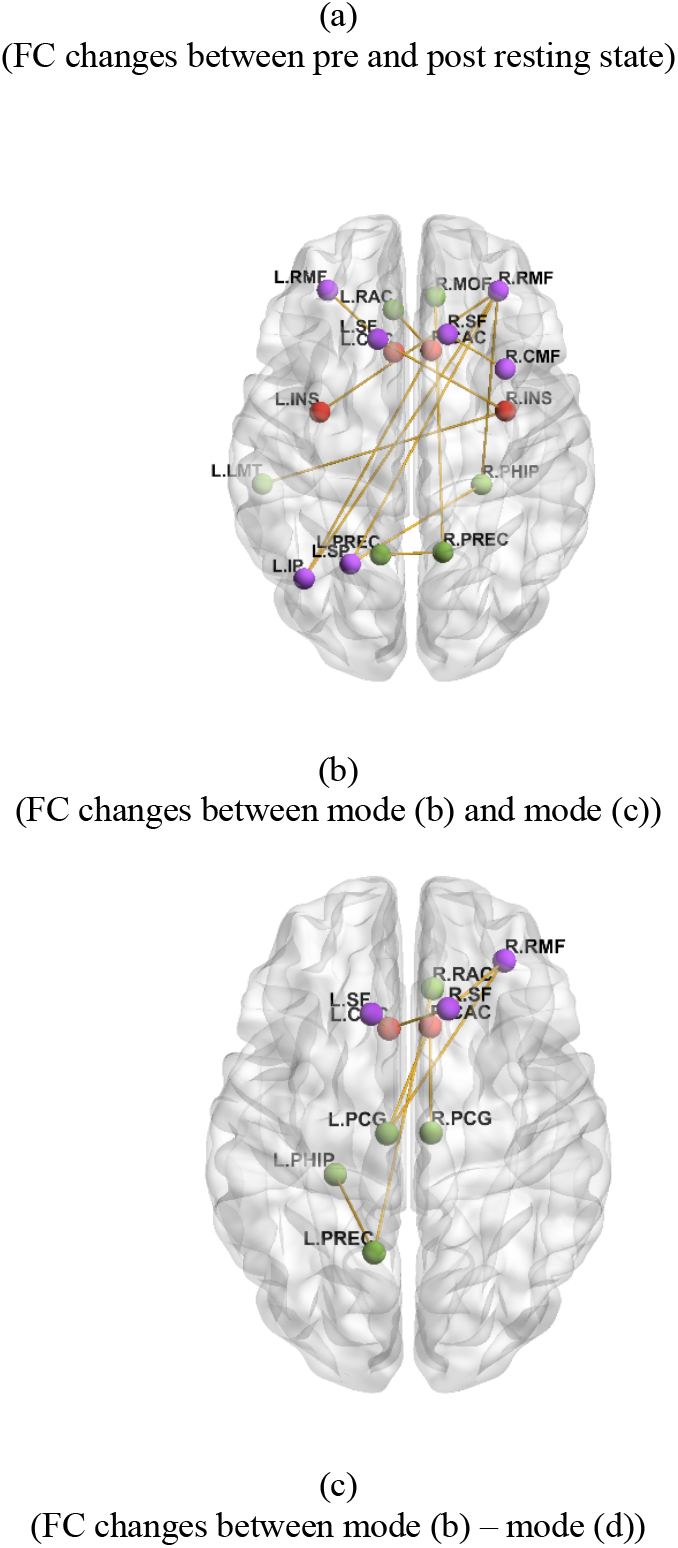
Significant changes in functional connectivity (PLI) in the beta band between (a) resting-state pre and post, (b) mode-(b) and mode-(c), and (c) mode-(b) and mode-(d). Spheres are cortical regions, coloured by network (1–3); lines are connections with significant differences (cluster-based permutation test, p < 0.05). Orange coloured lines indicates increase whereas blue decrease in the activity. Nodes are colored as: purple is CEN, green is DMN, and red is the Salience Network.

**TABLE 2.** Cutaneous sensation thresholds for healthy participants: (A) stochastic threshold, (B) sinusoidal vibration threshold, (C) sub-threshold combination, and (D) super-threshold combination. The final column reports the gain in sensitivity from mode (B) to mode(C).

| **Subject ID** | **mode (a)** | **mode (b)** | **mode (c)** | **mode (d)** | **Gain (b)→(c) (%)** |
|---|---|---|---|---|---|
| **1** | 0.3 | 16 | 10 | - | 60 |
| **2** | 0.4 | 11 | 2 | - | 450 |
| **3** | 0.2 | 8 | 5 | - | 60 |
| **4** | 0.3 | 10 | 1 | - | 900 |
| **5** | 0.3 | 5 | 1 | - | 400 |
| **6** | 0.3 | 2 | 1 | - | 100 |
| **7** | 0.4 | 7 | 4 | - | 75 |
| **8** | 0.3 | 3 | 1 | - | 200 |
| **9** | 0.3 | 3 | 1 | - | 200 |
| **10** | 0.3 | 2 | 1 | 0 | 100 |
| **11** | 0.3 | 3 | 1 | 0 | 200 |
| **12** | 0.9 | 12 | device minimum | 0 | - |
| **13** | 0.4 | 20 | 1 | 0 | 1900 |
| **14** | 0.4 | 7 | 4 | 0 | 75 |
| **15** | 0.4 | 10 | 1 | 0 | 900 |
| **16** | 0.3 | 2 | 1 | 0 | 100 |
| **17** | 0.3 | 2 | 1 | 0 | 100 |
| **18** | 0.3 | 2 | 1 | 0 | 100 |
| **19** | 0.4 | 20 | device minimum | 0 | - |

**TABLE 3.** Alpha-band region pairs with statistically significant changes in functional connectivity from cluster-based permutation analysis.

| Comparison | Region1 | Region2 | p value | RHO value |
|---|---|---|---|---|
| Resting state pre – post | R.IP | R.CMF | 0.0496 | 0.0059249 |
|  | L.MT | R.INS | 0.0376 | 0.0080911 |
|  | R.PHIP | L.CAC | 0.034 | 0.0099417 |
|  | R.PHIP | L.MOF | 0.0418 | 0.016603 |
|  | L.PREC | L.IP | 0.0322 | -0.0037062 |
|  | L.RAC | L.PCG | 0.0388 | 0.009546 |
|  | R.RAC | L.CMF | 0.0306 | 0.054797 |
|  | R.RAC | R.IP | 0.0164 | 0.0066662 |
|  | R.RAC | R.PHIP | 0.0344 | 0.014369 |
|  | L.RMF | L.CAC | 0.0114 | 0.040222 |
|  | L.RMF | R.CMF | 0.0216 | 0.041119 |
|  | L.RMF | L.MOF | 0.012 | 0.065314 |
|  | R.RMF | R.MT | 0.0352 | 0.0046778 |
|  | L.SF | R.RAC | 0.0468 | 0.048566 |
|  | R.SF | L.CMF | 0.0266 | 0.036616 |
|  | R.SF | R.INS | 0.0276 | 0.032524 |
|  | R.SF | L.RMF | 0.0234 | 0.053445 |
| mode (b) – mode (c) | R.IP | L.CAC | 0.0098 | 0.29441 |
|  | L.INS | L.CAC | 0.0414 | 0.2926 |
|  | R.INS | R.CAC | 0.0074 | 0.26579 |
|  | L.IST | L.CAC | 0.0062 | 0.26197 |
|  | L.MOF | R.IP | 0.0284 | 0.2461 |
|  | R.MOF | L.IP | 0.0278 | 0.24808 |
|  | R.MOF | R.INS | 0.0292 | 0.24664 |
|  | R.MT | R.CMF | 0.0396 | 0.2866 |
|  | L.PCG | L.PHIP | 0.0476 | 0.23969 |
|  | L.PREC | R.INS | 0.046 | 0.23427 |
|  | L.PREC | L.MOF | 0.0138 | 0.23998 |
|  | L.RAC | R.IP | 0.0108 | 0.24733 |
|  | L.RAC | L.PHIP | 0.0416 | 0.25604 |
|  | R.RAC | R.CAC | 0.0228 | 0.2532 |
|  | R.RAC | R.IP | 0.0188 | 0.2418 |
|  | R.RAC | R.PREC | 0.043 | 0.22924 |
|  | L.RMF | L.CMF | 0.004 | 0.29417 |
|  | L.SF | L.CAC | 0.0374 | 0.27331 |
|  | L.SF | L.INS | 0.0306 | 0.23928 |
|  | R.SF | L.IP | 0.0262 | 0.25989 |
|  | R.SF | R.INS | 0.0132 | 0.23608 |
|  | R.SF | L.MT | 0.0148 | 0.25332 |
|  | L.SP | R.IP | 0.0326 | 0.24929 |
|  | L.SP | R.RAC | 0.0038 | 0.22731 |
| mode (b) – mode (d) | L.RMF | L.PCG | 0.0172 | -0.0062004 |
|  | L.SF | L.MT | 0.0458 | -0.0054792 |

**TABLE 4.** Beta-band region pairs with statistically significant changes in functional connectivity from cluster-based permutation analysis.

| **Comparison** | **Region1** | **Region2** | **p value** | **RHO value** |
|---|---|---|---|---|
| Resting state<br>pre – post | R.CMF | L.CAC | 0.0132 | 0.008139 |
|  | R.IST | L.CMF | 0.038 | -0.00515 |
|  | L.MOF | L.INS | 0.0296 | 0.012947 |
|  | R.MOF | R.CAC | 0.0374 | 0.007387 |
|  | R.MT | R.MOF | 0.0232 | 0.006005 |
|  | R.PCG | R.CAC | 0.0272 | -0.00379 |
|  | R.PREC | R.IP | 0.0324 | 0.009536 |
|  | L.RAC | R.CAC | 0.0304 | 0.01174 |
|  | L.RMF | L.CMF | 0.0294 | 0.014569 |
|  | L.RMF | L.INS | 0.0202 | 0.013292 |
|  | L.SF | R.IP | 0.0448 | -0.00239 |
|  | R.SF | L.RAC | 0.0062 | 0.018341 |
|  | R.SF | L.RMF | 0.012 | 0.019357 |
|  | L.SP | L.PHIP | 0.0138 | 0.001603 |
|  | L.SP | L.RAC | 0.0316 | 0.003897 |
|  | L.SP | L.RMF | 0.0312 | 0.000962 |
|  | R.SP | L.INS | 0.048 | 0.00735 |
| mode (b) –<br>mode (c) | L.IP | R.CAC | 0.046 | 0.24936 |
|  | L.MT | R.INS | 0.0122 | 0.21405 |
|  | R.PREC | R.MOF | 0.0378 | 0.20798 |
|  | L.RAC | R.CAC | 0.0296 | 0.21354 |
|  | L.RMF | L.CAC | 0.0334 | 0.24346 |
|  | R.RMF | L.IP | 0.0228 | 0.2304 |
|  | R.RMF | L.INS | 0.0084 | 0.22419 |
|  | R.RMF | R.PHIP | 0.0418 | 0.21924 |
|  | L.SF | R.INS | 0.048 | 0.20447 |
|  | R.SF | R.CMF | 0.006 | 0.23381 |
|  | L.SP | R.PHIP | 0.0478 | 0.22579 |
|  | L.SP | R.RMF | 0.0466 | 0.21773 |
| mode (b) –<br>mode (d) | L.PCG | R.CAC | 0.0076 | 0.0074926 |
|  | R.PCG | R.CAC | 0.0414 | 0.0078632 |
|  | L.PCG | L.IP | 0.0124 | 0.0035259 |
|  | L.SF | L.CAC | 0.0372 | 0.0056795 |
|  | R.SF | L.CAC | 0.0258 | 0.011414 |
|  | R.MOF | R.INS | 0.046 | 0.00034057 |
|  | L.PREC | L.PHIP | 0.0422 | 0.0065359 |
|  | R.RMF | L.PCG | 0.0352 | 0.010222 |
|  | R.RAC | L.PREC | 0.0156 | 0.0082739 |
|  | R.RMF | R.SF | 0.0216 | 0.010422 |

## IV. Discussion

This study examined how sub-threshold stochastic vibration affects tactile detection thresholds and cortical FC in healthy adults and stroke survivors. Adding imperceptible stochastic noise to a sinusoidal stimulus lowered detection thresholds, that is, increased sensitivity, in every participant tested in healthy participants (individual sensitivity gains up to 1900%) and consistent reductions in all five stroke survivors (up to 400%). In parallel, source-localised EEG showed that sub-threshold vibration significantly increased cortical FC, most extensively in the alpha band. Together, these results indicate that sub-threshold stochastic vibration amplifies weak tactile signals and reorganises the cortical networks that process them.

These behavioural findings are consistent with, and extend, the established SR literature. Liu et al. reported that sub-threshold mechanical noise significantly lowered fingertip vibrotactile thresholds in older adults, in patients with diabetic neuropathy, and in four of five stroke patients, using the same 4–2–1 threshold-tracking procedure adopted here [5]; Enders et al. found that even remotely applied sub-threshold noise improved fingertip light-touch sensation in seven of ten stroke survivors, independent of location and intensity [23]. In the present cohort, all five stroke participants improved, and the direction and imperceptible-noise dependence of the effect reproduce these prior reports. Whereas earlier studies characterised the perceptual outcome and inferred a central contribution indirectly, the present study adds direct, source-localised evidence that the cortical networks supporting tactile processing are reorganised during sub-threshold stimulation. The connectivity results constrain the mechanistic interpretation. Under sub-threshold noise, alpha-band phase synchrony increased between superior frontal, rostral middle frontal, anterior cingulate, insular, inferior parietal and precuneal regions, and no significant pair in any contrast involved precentral or postcentral cortex. The regions involved are the nodes of the salience and frontoparietal control networks [24], [25]. Their state is known to determine whether a near-threshold somatosensory stimulus is perceived: baseline activity in this network predicts detection of an identical weak stimulus on a trial-by-trial basis [26], and pre-stimulus oscillatory activity over sensorimotor and parietal cortex influences detection of near-threshold tactile stimuli, while pre-stimulus alpha-band lateralisation over somatosensory cortex predicts tactile discrimination performance. [27], [28]. One account of alpha-band activity is that it gates sensory information by timed inhibition [29]. On that account, the present data are consistent with sub-threshold noise acting on the networks that gate perceptual access to a weak tactile signal, with the cortical representation of the signal itself unchanged at the resolution of this analysis. This reading fits the outcome measured, a lower detection threshold, and it fits the earlier finding that remote sub-threshold noise improves light-touch detection but not two-point discrimination [23]. It also fits the behavioural result, which is the expected signature of stochastic resonance in a threshold system [31], [32]. Because the phase lag index is insensitive to volume conduction [30], the increase in long-range synchrony reflects a change in inter-regional coupling rather than the spread of a single source. The same measure discards zero-lag coupling [30], so a zero-lag interaction between primary somatosensory and motor cortex would not have been detected, and the absence of a primary sensorimotor pair is not evidence that primary sensorimotor cortex is unaffected. The design is correlational and used one sub-threshold noise level, so whether the change in alpha-band phase consistency contributes to the lower threshold or accompanies the attentional state that produces it cannot be decided from these data.

The contrast between sub- and super-threshold conditions reinforces the SR interpretation. Consistent with Collins et al., who showed that noise enhances tactile detection only below the perceptual threshold and degrades it above [4] (as also emphasised by Enders et al. [23]), the 120% super-threshold noise did not confer a sub-threshold–type benefit: it was itself always perceptible (thresholds recorded as 0) and was accompanied by markedly weaker, predominantly beta-band FC changes, with minimal alpha-band involvement (two region pairs). In other words, once the added noise is supra-threshold it masks rather than resonantly amplifies the target signal, engaging a different, and less extensive, connectivity pattern than sub-threshold stimulation. The stronger and more widespread FC changes under sub-threshold [mode (c)] than super-threshold [mode (d)] stimulation suggest that imperceptible noise more effectively engages latent sensory pathways. Again, understanding the detailed mechanistic action requires future studies.

For stroke survivors, the immediate improvement in tactile thresholds, reproducing Liu et al. [5] and Enders et al. [23] in an independent cohort, supports sub-threshold stochastic vibration as a non-invasive route to enhancing impaired cutaneous sensation. Because cutaneous feedback is essential to grip control and dexterity [23], restoring light-touch sensitivity could, in principle, support sensorimotor recovery, although this requires direct investigation. The accompanying FC changes are consistent with a capacity of stochastic noise to promote cortical reorganisation, a key substrate of post-stroke recovery.

Several limitations should be noted. First, the stroke cohort was small (n = 5) and the super-threshold condition was limited to ten healthy participants, constraining the generalisability of the FC findings; larger, adequately powered cohorts are needed. Second, individual responses varied widely (sensitivity gains from 16% to 1900%), pointing to person-specific determinants of SR efficacy—such as age, baseline sensitivity, lesion characteristics, and neural plasticity—that warrant systematic study. Third, although we observed both threshold reductions and FC increases, we did not compute a formal brain–behaviour correlation between the two; establishing whether the magnitude of FC change predicts the magnitude of perceptual improvement is an important next step (cf. the correlation analyses used in prior work [23]). Fourth, we assessed detection thresholds only; Enders et al. found that sub-threshold noise improved light-touch detection but not two-point (spatial) discrimination [23], so future studies should include spatial-acuity and functional (dexterity, grip-force) measures to determine which aspects of sensation and hand function are enhanced. Finally, we measured immediate effects; the durability of both perceptual and connectivity changes, and their response to repeated or longer-term exposure, remain to be established.

## V. Conclusion

Sub-threshold stochastic vibration significantly enhanced tactile sensitivity and reorganised cortical FC in both healthy adults and stroke survivors. Combining imperceptible stochastic noise with sinusoidal stimulation lowered detection thresholds in every participant in healthy participants (individual sensitivity gains up to 1900%) and in all five stroke survivors (up to 400%), consistent with stochastic resonance amplifying weak sensory signals. Source-localised EEG revealed frequency-specific network reorganisation, with the most extensive increases in alpha-band connectivity during sub-threshold stimulation and a shift toward beta-band involvement under super-threshold stimulation, the latter consistent with signal masking rather than resonance.

By pairing psychophysical thresholds with direct, source-level connectivity evidence, this work links noise-enhanced perception to measurable cortical reorganisation, extending prior behavioural demonstrations of somatosensory stochastic resonance [5], [23] toward a mechanistic account. The main limitations are the small stroke cohort (n = 5) and the wide inter-individual variability in response. Future work should pursue longitudinal designs, individualised noise parameters, formal brain–behaviour correlations, and combined sensory–motor outcomes, with the goal of translating sub-threshold stochastic vibration into practical, non-invasive sensory rehabilitation after stroke.

## Data Availability

All data produced in the present study are available upon reasonable request to the authors

## Acknowledgment

Authors sincerely thank all the participants for taking part in this study.

